# Comprehensive genomic atlas of plasma proteome in the Japanese population: the Nagahama study

**DOI:** 10.1101/2025.11.20.25340712

**Authors:** Ta-Yu Yang, Takahisa Kawaguchi, Shuji Kawaguchi, Koichiro Higasa, Satoshi Yoshiji, Takeshi Iwasaki, Tie Zhao, Meiko Takahashi, Kazuya Setoh, Katsunori Horii, Shintaro Kato, Takeo Nakayama, Shinji Kosugi, Iwao Waga, Yasuharu Tabara, Masao Nagasaki, Fumihiko Matsuda

## Abstract

Combining the plasma proteome with the genome offers insights into diseases. Here, we characterized the genetic architecture of plasma proteins in 1,823 Japanese individuals. We identified 1,876 protein quantitative trait loci (pQTLs) comprising 1,395 variants associated with 1,254 unique proteins, with 77 pQTLs being specific to the Japanese population. Multi-ancestry fine-mapping identified 475 credible sets shared between Japanese *cis*-pQTLs and European *cis*-eQTLs. By integrating both *cis*- and *trans*-pQTLs, we identified a Japanese-specific *trans*-pQTL hotspot in the *CD36* gene, associated with 10 proteins and linked to decreased platelet and white blood cell counts. Leveraging Mendelian randomization (MR) integrating pQTLs and Biobank Japan genome-wide association study (GWAS), we identified 42 putative causal relationships between 24 proteins and 20 diseases. This analysis identified 11 proteins (FCRL1, KLB, ADH1B, ADH1C, IL1RL1, IL18R1, DPEP1, HP, MICB, IL6R, LRP11) as potential drug targets. Our findings significantly enhance the understanding of the plasma proteomic landscape in an East Asian population and provide a valuable resource for prioritizing population-specific therapeutic targets.

## Introduction

Proteins play a pivotal role in various biological processes, such as host defense, signaling, and cellular metabolism^1^. Genetic variation can alter the abundance and functions of proteins, which may be associated with diseases. Understanding the association between genetic variants and plasma protein levels, as assessed by high-throughput measurements of circulating proteins, facilitates the elucidation of disease etiology and the identification of potential druggable targets^1^. With the rapid advances in aptamer-and antibody-based protein quantification technologies^2,3^, several studies have analyzed protein quantitative trait loci (pQTL) to broaden the current understanding of genomic variants influencing plasma protein levels and disease pathways^4–9^. However, because most pQTL studies have focused predominantly on European populations^6,8,10,11^, there is a strong rationale for incorporating other populations, given the impact of differences in genomic and environmental factors on phenotypes among populations. Using a population-specific protein dataset facilitates exploring of the mechanisms through which genomic variants affect plasma protein levels^4,12^. By conducting a two-sample MR analysis integrating pQTLs and GWAS on various disease phenotypes within the Japanese population, we may be able to detect potential causal proteins and actionable drug targets. This approach not only contributes to the identification of relevant proteins but also enriches our insights into cross-population proteomic analyses.

In this study, we leveraged Nagahama cohort, a Japanese cohort comprising 1,823 individuals with whole-genome sequencing (WGS) data and 4,239 measured plasma proteins to conduct pQTL analysis. Furthermore, we evaluated causal relationships between plasma protein levels and diseases and identified potential causal proteins and druggable targets using MR that combined pQTL data with 65 disease phenotypes obtained from BioBank Japan. This comprehensive analysis provides valuable insights into the complex relationships between the plasma proteome and diseases.

## Methods

### Study cohort

Whole-genome sequencing and proteomic analysis were conducted using biomaterials collected from participants in the Nagahama Prospective Genome Cohort for Comprehensive Human Bioscience (the Nagahama Study)^13^. A total of 2,000 individuals, including 1,392 women (mean age 56.7 years) and 608 men (mean age 62.0 years), were selected from 8,559 participants who participated in the first follow-up health examination between 2012 and 2016, and their DNA and plasma samples were used for the analysis. All procedures were approved by the ethics committees of Kyoto University Graduate School of Medicine and the Nagahama Municipal Review Board (No. 278). Participants were fully informed of the purpose and procedures of this study, and all participants provided written informed consent.

### Plasma samples and protein quantification

Plasma was isolated by centrifugation at 3000 rpm at 4 °C for 15 min from EDTA-treated blood specimens drawn during the first follow-up. The plasma samples were stored at−80 °C until plasma protein analysis. Plasma protein levels were quantified using the SomaScan version 4 assay (SomaLogic, Inc., Boulder, CO, USA) with 5,284 slow off-rate modified aptamers (SOMAmers) corresponding to 4,740 unique proteins (**Supplementary Table 1**).

Protein quantification was conducted at SomaLogic Inc. The results were returned after quality control according to the standard protocol of SomaLogic (SL00000442, SM-00060-Rev1.0). Three plasma samples and 526 SOMAmers were removed during this step owing to quality issues. Additionally, 305 SOMAmers labeled by SomaLogic, including non-human, spuriomer, hybridization control elution, deprecated, non-biotin, and non-cleavable SOMAmers, were excluded. Gene and protein sequences were annotated using the Ensembl database. After annotation, we removed 61 SOMAmers, including those with gene annotation withdrawn by NCBI or Ensembl (seven SOMAmers) and those targeting protein complexes with at least two subunits corresponding to different genes (39 SOMAmers). Furthermore, SOMAmers targeting HSP70 (8) and tenascin (7) were excluded owing to unknown functions in multiple SOMAmers. Finally, 1,997 plasma samples containing 4,392 SOMAmers (4,196 proteins) passed the quality control steps, and inverse normalization was performed. The normalized data were used in the pQTL analysis. In our study, most analyses were based on the measurement of SOMAmers; however, some SOMAmers were designed to target the same protein in different regions or forms. In the SomaScan dataset, they can be distinguished by annotation in the SomaId and Target columns.

### Whole-genome sequencing

Among the 1,997 samples, WGS was performed using the Illumina HiSeq X (n = 1,232), NovaSeq 6000 (n = 259), and HiSeq 2500 (n = 82) platforms, and 424 samples did not have WGS data. These 424 samples were first checked through kinship analysis using a single nucleotide polymorphism (SNP) array, and 39 were excluded from the study. WGS was then performed on the remaining 385 samples using a DNBSEQ-G400 instrument (MGI Tech Co., Ltd., China). The Genome Analysis Toolkit (GATK) variant-calling pipeline (version 4.1.4.0)^14^ was executed on the 1,573 Illumina WGS samples, and the DRAGEN germline small variant caller (version 3.8) with default settings^15^ was used on the 385 samples from the DNBSEQ analysis. The GRCh38 reference genome was used in both the GATK and DRAGEN pipelines.

After obtaining the variant call format files (VCFs) from both the GATK and DRAGEN pipelines, we assessed variant concordance between the SNP array and WGS using the SNP array results from the same samples. In this step, five additional samples from Illumina and seven samples from DNBSEQ that showed discordance were removed. Two further Illumina samples and one DNBSEQ sample were also excluded. Finally, the VCF produced by GATK containing 1,566 samples from Illumina WGS and the VCF containing 377 samples produced by the DNBSEQ and DRAGEN pipelines were merged. To combine the two VCFs, variant-and sample-level quality control for the VCF from each pipeline were performed. First, after obtaining unfiltered variants from the VCF, variants that contained only the word “PASS” in the FILTER column were selected. Second, we applied additional filters for DRAGEN in the GT column to convert the genotype to “no-call (. /.)” in cases where GQ < 20, DP < 5, or allele balance < 0.2 or > 0.8. Variants were then selected where AC ≥ 1. For the VCF generated through GATK, genotypes satisfying the filter threshold DP < 10, and the allele balance threshold ≤ 0.25 were changed to no-call within the GT column. Two additional filters, variant quality score log odds > 10 and mapping quality > 58.75, were applied to the VCF from the GATK pipeline. After merging the two VCFs, variants with call rates < 0.9 and Hardy-Weinberg equilibrium *P*-values < 1 × 10^−6^ were removed. Finally, variants with a minor allele frequency < 0.01 were removed. For sample quality control, 16 samples with a call rate of < 0.8 and 104 samples showing cryptic relatedness (PIHAT > 0.15) were removed. After the quality control steps, we obtained 1,823 samples and 4,642,253 variants that were ready for protein association analysis (**Supplementary Table 2**).

### Variant-protein association and conditional analysis

We analyzed associations between genetic variants and 4,392 SOMAmers using PLINK (version 2.00a3LM)^16^ with the --glm function. The covariates included in the association model were age, sex, variant-calling pipeline, and the first five principal components. To identify conditional independent signals, we performed conditional analysis using GCTA-COJO^17^ with the following parameters: “--maf 0.01 --cojo-slct --cojo-collinear 0.9 --cojo-p 5e-8.” The multiple-testing significance threshold (1.14 × 10^−11^) was determined using genome-wide significance divided by the number of tested SOMAmers (5 × 10^−8^ / 4392).

Variant-protein associations were classified as “*cis*” when the variant was within the gene region, defined as the protein-coding gene (annotated using Ensembl Variant Effect Predictor (VEP) version 110) extended by 500 kb upstream of the gene start position and 500 kb downstream of the gene end position (**Supplementary Table 3**). The variant-protein associations were defined as “*trans*” when the variant was located outside the ± 500 kb defined *cis* gene. The lead *cis*-acting variant associated with a given SOMAmer was assigned as the one with the lowest conditional *P*-value, calculated using GCTA-COJO. The lead *trans*-acting variant was assigned using the same approach.

### Explained genetic variance

The explained genetic variance (*R^2^*) of each SOMAmer was estimated using the equation described by Park et al^18^. In summary, we used beta and SE estimates generated by GCTA-COJO, as well as the effect allele frequency, to calculate the explained genetic variance for each independent variant.

### Reported associations in European and African American population

To assess shared pQTLs between the Japanese population and other populations using SomaScan platform, we retrieved summary statistics from a European study (deCODE^8^; 4,907 SOMAmers) and an African American study (ARIC^12^; 4,657 SOMAmers). We restricted our analysis to associations reaching genome-wide significance (*P* = 5 × 10^−8^).

Notably, the publicly available ARIC dataset contained only *cis*-pQTLs. Following the harmonization of SOMAmers, we defined a shared pQTL as the presence of a significant signal within a 500 kb window of the corresponding pQTL in the comparator dataset.

### Linkage disequilibrium (LD) clumping

For each chromosome, we gathered the associated variants and *P*-values for each pQTL and performed LD clumping. The variant with the lowest *P*-value was selected as the index variant. A clumping region was defined as 500 kb around the index variant, and the LD threshold was set to be *R*^2^ ≥ 0.8. PLINK (version 1.9) was used with the following parameters: “--clump-kb 500 --clump-r2 0.8 --clump-p1 1 --clump-p2 1”. After obtaining the results, each LD clumping region was assigned a unique identifier.

### Annotation of variants

Variants were annotated using the Ensembl Variant Effect Predictor (VEP; version 110)^19^. According to the VEP annotation, HIGH-impact variants included splice acceptor, splice donor, stop-gained, frameshift, stop-lost, and start-lost. MODERATE-impact variants included missense variants, in-frame insertions, and in-frame deletions. The subcellular location of the protein was determined by searching UniProtKB^20^ on July 13^th^, 2023. The effect allele frequency of the variants was annotated using 1,000 Genomes Project Phase 3 and gnomAD genomes version 3.1.2.

### Protein-altering variants (PAVs)

PAVs were defined as HIGH-impact or MODERATE-impact variants using VEP annotation. To analyze the possibility of pQTLs being affected by PAVs, we calculated LD between each pQTL and PAVs within the locus. If a pQTL was in high LD (*R^2^* > 0.8) with a PAV, they were tagged as potential PAV candidates.

### Multi-ancestry fine mapping and colocalization analysis

The expression quantitative trait loci (eQTL) datasets were downloaded from GTEx (version 8). The Ensembl gene ID was used to match the annotation between the measured proteins and genes in GTEx. Fine mapping was performed using the R package MESuSiE^21^. Variants with a posterior inclusion probability (PIP) ≥ 0.5 were considered shared causal variants between European and Japanese populations. Furthermore, a *P*-value < 1 × 10^−5^ was used to filter out non-significant causal variants from *cis*-eQTL signals, as suggested by GTEx^22^. For the Japanese-specific gene expression dataset, we utilized the publicly available *cis*-eQTL summary statistics^23^. Variants that passed QC (i.e., sample missing call rate ≤ 0.01, minor allele frequency ≥ 0.05, and Hardy-Weinberg equilibrium *P*-value ≥ 1e-7) were selected, and only variants with a corresponding rsID were retained. By matching rsID with dbSNP 154, we converted the genomic position of GRCh37 to GRCh38. Ultimately, we identified 768 genes with a total of 318,404 variants. Colocalization analysis was performed using the coloc R package^24^ (version 5.0.1). The posterior probability of hypothesis H4 (PP.H4) ≥ 0.8 was used to identify shared causal variants between proteins and gene expression.

### Protein-protein interactions and pathway enrichment analyses

The STRING (version 12^25^ was used to analyze protein-protein interactions and to perform pathway enrichment analysis. Default settings were used for network analysis, and the Entrez Gene Symbols of the selected SOMAmers were used as input to construct the network.

### Mendelian randomization analysis and drug target evaluation

From the BioBank Japan PheWeb^26^, data on 65 diseases based on the International Classification of Diseases, 10th Revision (ICD-10), were collected, each with more than 1,000 cases (**Supplementary Table 7**). We used conditionally independent variants obtained from the previous step as instrumental variables (IVs), which were checked to meet three MR assumptions^27^: (i) Relevance: genetic variants are associated with the exposure of interest, (ii) Independence: genetic variants share no unmeasured causes with the outcome, and (iii)

Exclusion restriction: genetic variants do not affect the outcome except through their potential effect on the exposure of interest. We retained protein exposures with an F-statistic > 10^28^. MR Steiger filtering^29^ was performed to confirm the direction and causal effects (Steiger *P*-value < 0.05). We removed IVs classified as PAVs or in LD with PAV (*r^2^* > 0.8), unless they were identified as significant eQTLs (*P* < 1 × 10^−5^). To examine horizontal pleiotropy, we used MR-egger^30,31^ for results containing multiple IVs. When only one IV was available for an exposure, we used PhenoScannerV2^32^, Open Target Genetics^33^, and the GWAS Catalog^34^ to evaluate potential pleiotropic effects. To identify potential drug targets, we queried protein targets using ChEMBL^35^ (https://www.ebi.ac.uk/chembl/, release 35) through two approaches:

(i) ID mappings by querying Ensembl gene IDs and (ii) utilizing sequence similarity nearest neighbors based on UNIREF clusters^36,37^.

## Results

### Associations between plasma protein levels and genetic variation in the Japanese population

We performed variant-protein association analyses involving 4,642,253 variants and 4,392 slow-off-rate modified aptamers (SOMAmers), corresponding to 4,239 unique proteins. Because multiple SOMAmers can target the same protein and may bind different domains, they can provide distinct information. Thus, throughout the manuscript, we use the term “SOMAmers” to refer to proteins unless otherwise specified. Detailed sample characteristics can be found in the published Nagahama cohort profile^13^ and **Supplementary Table 1**, and protein information is shown in **Supplementary Table 2**. We tested variant-protein associations and identified 1,876 conditionally independent associations (pQTLs) that passed a multiple testing-corrected threshold of *P* < 1.14 × 10^−11^ (5 × 10^−8^ / 4392), comprising 1,275 SOMAmers corresponding to 1,243 proteins based on UniProt ID and 1,395 variants.

Genomic inflation was well controlled, with a median λ_GC_ = 1.005 (s.d. = 0.007). Of the 1,275 SOMAmers, 855 (67%) were associated with a single variant, and 273 (21%) were associated with two variants (**Fig. 1a**). Our results showed that 1,226 (88%) of variants were associated with only one SOMAmer, while 169 variants (12%) were associated with at least two SOMAmers. Of those 169 variants, the maximum number of SOMAmers associated per variant was 24. Among the 1,876 pQTLs, 936 were *cis*-pQTLs and 940 were *trans*-pQTLs (**Supplementary Table 3**). Furthermore, we identified 109 SOMAmers that had both *cis* and *trans* associations, 570 SOMAmers were associated with only *cis*-acting variants, and 596 SOMAmers were associated with only *trans*-acting variants.

**Fig. 1a.**
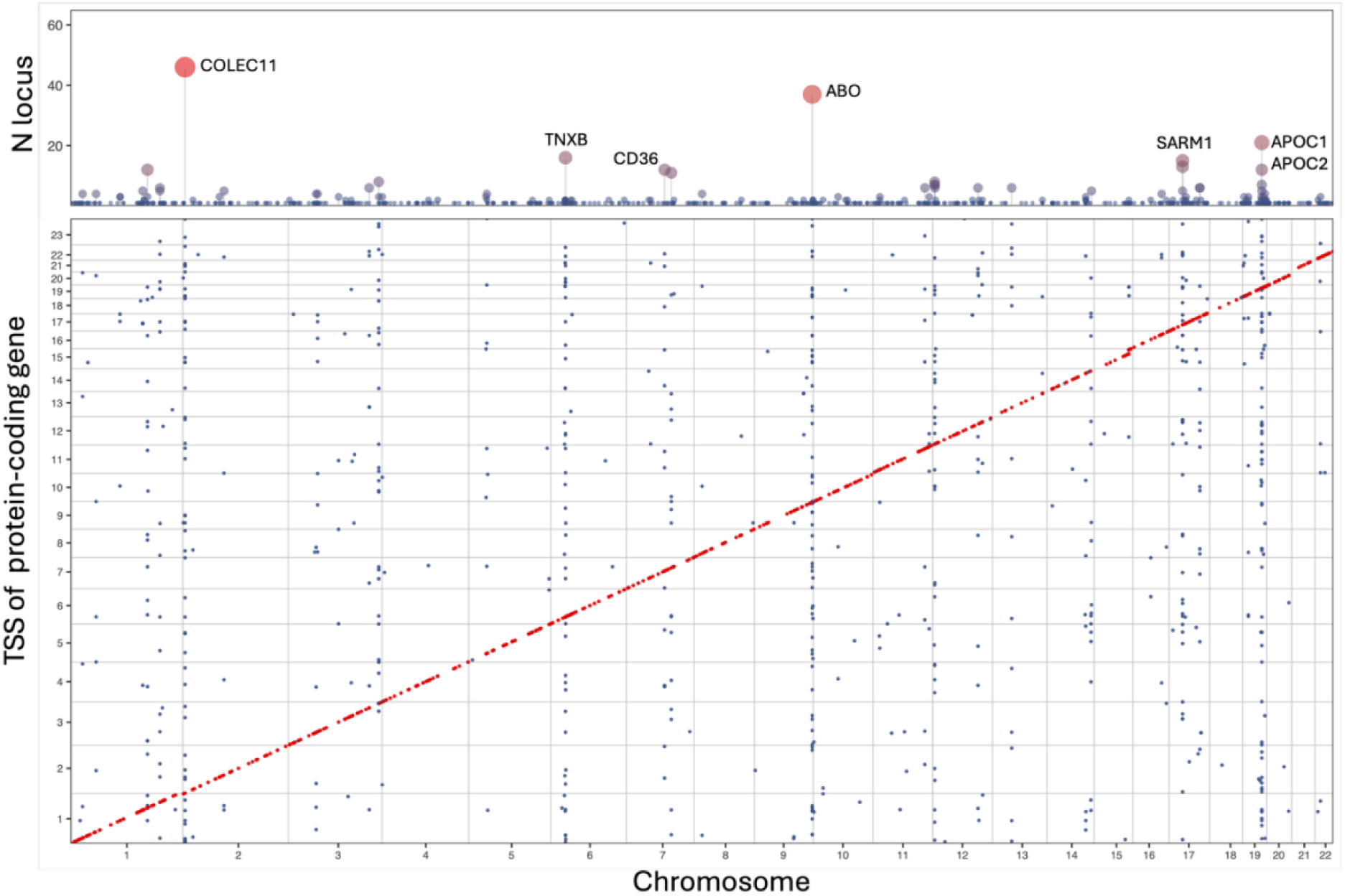
Overview of protein quantitative trait loci (pQTLs) in the Japanese population. Genomic locations of 1,384 lead pQTLs (bottom) colored according to *cis*- (red) and *trans*-acting (blue) variants. The x-axis indicates the genomic position of the lead variants, and the y-axis indicates the transcription start site (TSS) of the corresponding protein-coding gene. Numbers of LD-clumped loci are plotted in the top panel. The y-axis indicates the number of loci.

**Fig. 1b.**
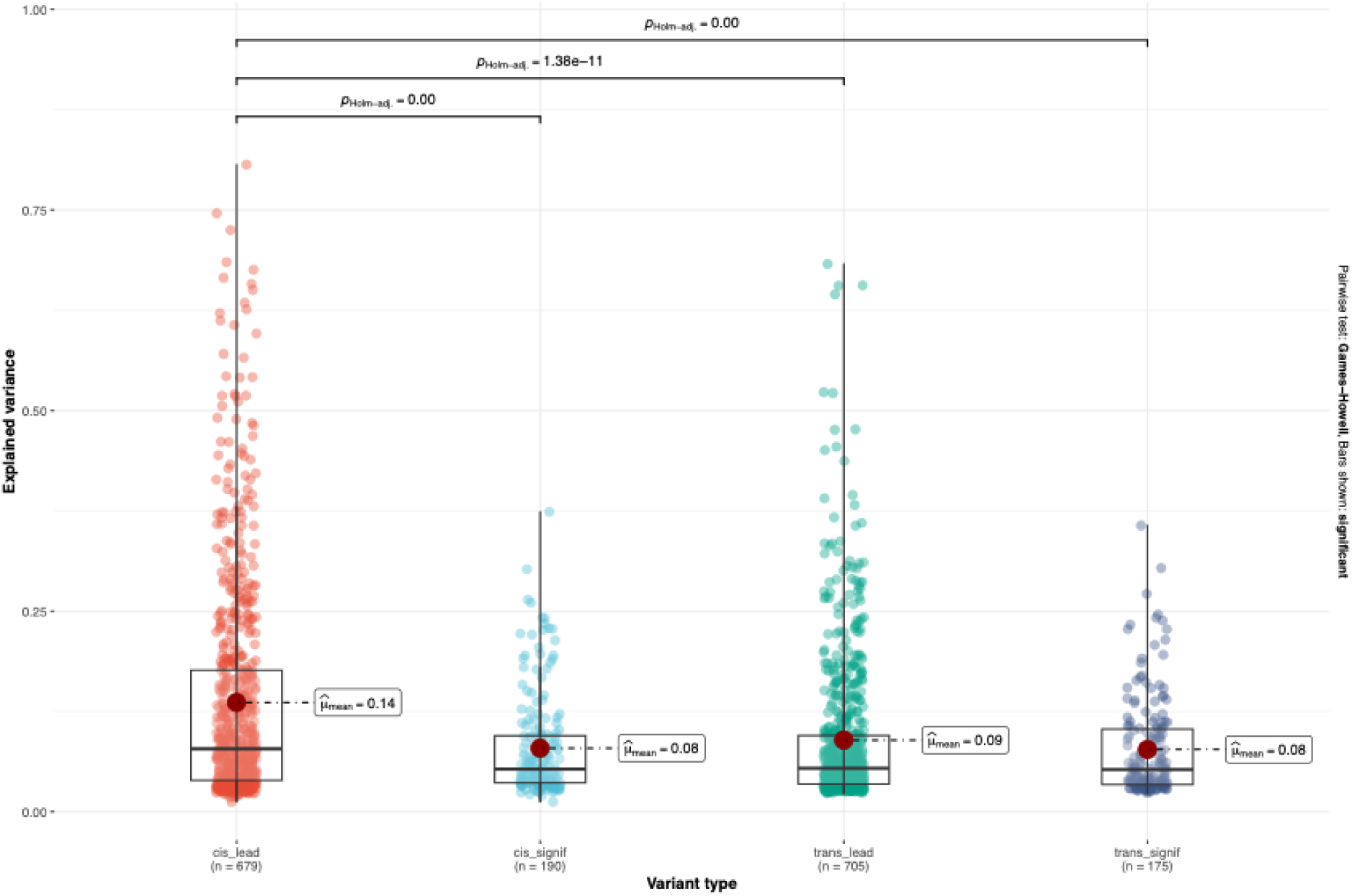
Explained variance across different types of pQTLs. Box plot of the proportion of variance in plasma protein levels explained by different types of pQTLs. The y-axis indicates the explained variance (*R^2^*) value. The x-axis indicates the type of pQTL used for variance calculation.

### Genetic variance in plasma protein levels

To estimate the contribution of genetic variants to the variance of each plasma protein level, a lead association was first assigned based on the variant with the lowest *P*-value associated with that protein after performing GCTA-COJO. Other conditionally independent variants associated with the same protein were then marked as significant associations. The genetic variance in plasma protein levels was explained using two approaches for each protein: (1) the lead association only and (2) all associations excluding the lead association. For lead *cis*-acting variants, the mean variance explained was 13.6%. All conditionally independent *cis*-acting variants, excluding lead variants, collectively explained a mean variance of 7.9%. Similarly, lead *trans*-acting variants explained a mean variance of 8.9%, and all conditionally independent *trans*-acting variants, excluding lead variants, showed a mean variance of 7.9% (2.4–8.8%) (**Fig. 1b**). These findings indicate that a greater proportion of the genetic variance in plasma protein levels is accounted for by *cis*-acting variants, particularly when lead *cis*-acting variants are included.

### Genetic architecture of pQTLs across diverse ancestries

To investigate the genetic architecture of pQTLs between the Japanese population and other populations, we compared our results with a European study (deCODE) and an African American study (ARIC) using summary statistics from each cohort, restricting analyses to associations that reached genome-wide significance (*P* = 5 × 10^−8^). To assess trans-ancestry concordance, we compared our pQTLs with those reported in the deCODE (European) and ARIC (African American) datasets after harmonizing SOMAmers. We defined a shared pQTL as a signal identified within 500 kb of a pQTL in the comparator dataset. We found high concordance between the European and Japanese populations, with 1,796 of 1,876 (96%) pQTLs shared, which showed strongly correlated effect sizes (r_Pearson_ = 0.90, 95% CI: 0.89–0.91, *P* = 2.2 × 10^−16^; **Supplementary Table 3; Supplementary Figure 1**). The comparison between the African American and Japanese populations identified 935 shared pQTLs (50%), which also demonstrated highly correlated effect sizes (r_Pearson_ = 0.89, 95% CI: 0.87–0.91, *P* = 1.1 × 10^−153^; **Supplementary Figure 2**). Notably, these shared signals were exclusively *cis*-pQTLs, as the publicly available African American dataset was restricted to *cis*-variants. We also observed 932 pQTLs (50%) that were shared across the three populations. Furthermore, we identified 77 pQTLs (4%) that were not reported in either the deCODE or ARIC studies.

### Population-specific pQTLs in the Japanese population

When comparing the effect allele frequencies (EAF) of 77 pQTLs unique to the Japanese population with those of Europeans and African Americans, the results indicated a higher EAF among the newly identified Japanese-specific pQTLs (**Fig. 2**). Of the 77 pQTLs, 43 had an EAF > 0.01 in Japanese but an EAF < 0.01 or were absent in European populations.

**Fig. 2.**
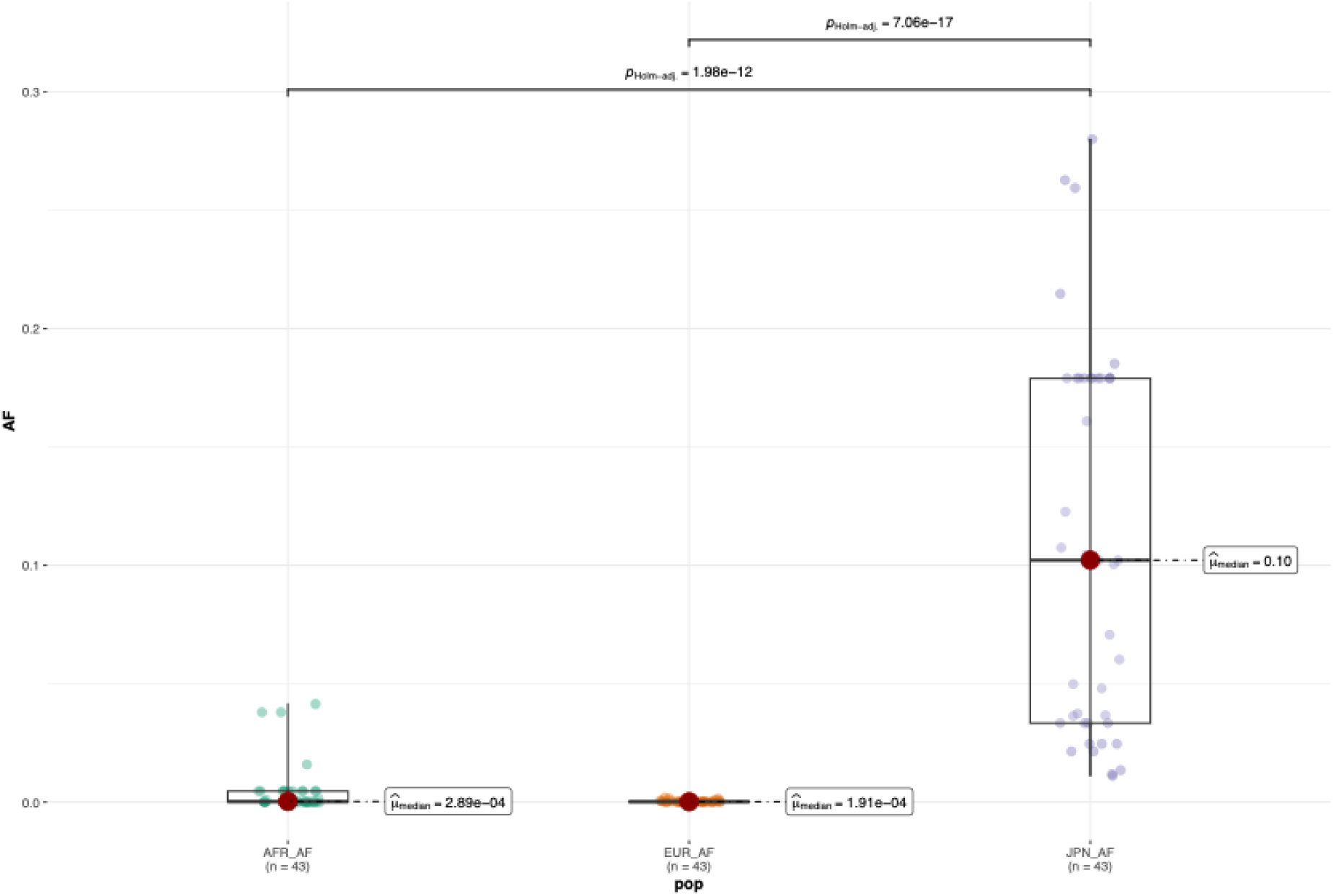
Differences in effect allele frequency of Japanese-specific pQTLs across populations. Boxplots of the effect allele frequency for 77 Japanese-specific pQTL in three populations. The x-axis shows populations (AFR, African; EUR, European; JPN, Japanese), and the y-axis shows alternative allele frequency.

Among these 43 pQTLs, we identified 42 *trans*-pQTLs and one *cis*-pQTL. The *cis*-acting variant (rs150785815T>C, effect allele: C) was associated with Kit ligand (SCF) protein, and the effect allele decreased plasma SCF protein levels (beta =-0.88, *P* = 5.89 × 10^−23^). This variant had an allele frequency of 0.03 in our cohort. However, the allele frequency in the European population was virtually zero (EAF < 4.4 × 10^−5^). Furthermore, of the 42 identified *trans*-pQTLs, 12 proteins were associated with two variants (chr7:80569431:C:T, chr7:80574911:GC:G), which had very low allele frequencies in European (EAF = 0.0003) compared with Japanese populations (EAF = 0.18). By searching these two variants on the BioBank Japan PheWeb, we identified that both variants were associated with platelet count (beta =-0.022, *P* = 1.4 × 10⁻^7^) and white blood cell count (beta =-0.022, *P* = 7.0 × 10⁻^7^). The two variants were located in the *CD36* gene and are in high LD (*r^2^* = 0.94), suggesting a population-specific pleiotropic effect at this locus. These results highlight the influence of population-specific variants, which could alter the plasma proteome.

### Integrating *cis*-eQTL in fine-mapping analysis

To discern the causal variants influencing plasma protein levels, we leveraged gene expression datasets from GTEx version 8. Since the GTEx participants are predominantly of European ancestry, we used MESuSiE, a multi-ancestry fine-mapping approach (see **Methods**), combining our *cis*-pQTLs and *cis*-eQTLs from GTEx. Our pQTLs identified 660 proteins associated with *cis*-acting variants, with 656 of these having protein-coding genes listed in the GTEx database. We identified 411 out of 656 genes (63%) that were expressed specifically in a given tissue, as defined by the tissue-specificity metric tau (𝜏) > 0.85. As over 60% of protein-coding genes were highly expressed in specific tissues, we therefore selected the tissue with the highest gene expression for each gene to perform multi-ancestry fine-mapping. Out of 656 proteins, we identified 173 SOMAmers (26%; 170 proteins) that were associated with shared credible sets in both Japanese and European populations, comprising 475 shared *cis*-pQTLs and *cis*-eQTLs, respectively. The correlation between the effect sizes of shared variants was moderate (r_Pearson_ = 0.57, 95% CI: 0.51–0.63, *P* = 2.1 × 10^−42^; **Supplementary Figure 3).**

To investigate the correlations between gene expression and plasma protein levels, proteins were labeled with their subcellular location using the UniProt database. Out of 173 SOMAmers, 125 SOMAmers (123 proteins) had labels indicating they were either “Secreted” (79), “Cell Surface” (2), “Cell membrane” (38), or “Membrane” (17), where some SOMAmers had multiple annotations. The remaining 48 SOMAmers (48 proteins) were located within the cell. After categorizing SOMAmers by their subcellular locations, we observed that the correlation of effect size was higher in the 327 pQTLs associated with 125 secreted or membrane SOMAmers (r_Pearson_ = 0.59, 95% CI: 0.51–0.65, *P* = 7.78 × 10^−32^; **Supplementary Figure 4**) compared with those 148 pQTLs associated with 48 SOMAmers (r_Pearson_ = 0.56, 95% CI: 0.43–0.66, *P* = 2.23 × 10^−13^; **Supplementary Figure 5**). For 125 secreted or membrane SOMAmers, we stratified them based on their associations with protein-altering variants (PAVs) and non-PAVs (see **Methods**). Among 327 *cis*-pQTLs, 36 variants were protein-altering variants (PAVs) or were in high LD (*r^2^* > 0.8) with PAVs. No significant correlation was observed between the effect sizes of these 36 *cis*-pQTLs and *cis*-eQTLs (r_Pearson_ = 0.31, 95% CI:-0.02–0.58, *P* = 0.06; **Supplementary Figure 6**). In contrast, a higher correlation was evident for the remaining 291 non-PAV *cis*-pQTLs (r_Pearson_ = 0.65, 95% CI: 0.58–0.71, *P* = 4.98 × 10^−36^; **Supplementary Figure 7**). We also identified 19 of 173 SOMAmers associated with 18 Japanese-specific credible sets; these sets were distinct from those shared with European populations. A subsequent comparison between the effect sizes of these 18 Japanese-specific sets and GTEx *cis*-eQTLs showed no correlation (r_Pearson_ = 0.37, 95% CI:-0.04–0.67, *P* = 0.08).

In addition to the GTEx eQTL dataset, we performed colocalization analysis using the Nagahama *cis*-eQTL dataset, generated from whole blood and comprising 768 gene expression profiles and 318,403 variants (see **Methods**). After harmonizing genes between eQTL and pQTL, we conducted colocalization analysis based on 679 lead *cis*-pQTL (see **Methods**). We identified 72 of 679 (11%) *cis*-pQTLs colocalized with a *cis*-eQTL (*r*_Pearson_ = 0.68, 95% CI: 0.53–0.79, *P* = 6.96 × 10^−11^; **Supplementary Figure 8**). By integrating MESuSiE and colocalization analyses, we identified 173 SOMAmers shared between the two populations and 72 SOMAmers with Nagahama eQTLs, respectively.

### Elucidating the pathway using *cis*- and *trans*-pQTLs

To elucidate the relationship between *cis* and *trans* associations and their combined effect on protein pathways, we defined 876 loci using LD clumping (**Fig 1a**, top; see **Methods**). For each locus, the variant with the lowest *P*-value was assigned as the index variant. Four loci were associated with more than 20 SOMAmers, and eight were associated with at least 10 SOMAmers (**Supplementary Table 4**). Among the 876 loci, there were 603 *cis*-only loci, 225 *trans*-only loci, and 48 loci containing both *cis* and *trans* associations. One locus (designated as chr2_L6) near the *COLEC11* gene, with index variant (chr2:3588888:T:G), was associated with 46 SOMAmers corresponding to 46 proteins (**Supplementary Table 5**), with one *cis-* and 45 *trans*-pQTLs. The variant rs56236159T>G was in strong LD with rs3820897, one of the lead variants (chr2:3594771:T:C, *r^2^* = 0.976) within the promoter region; it reportedly affects the abundance of collectin-11 (COLEC11)^38^. We performed pathway and protein-protein interaction (PPI) analyses for these 46 proteins (see **Methods**). Our results showed that IL1B and COLEC11 interacted with 19 other proteins with a PPI enrichment *P*-value of 1.64 × 10^−4^ (**Fig. 3a**). Furthermore, the cell surface receptor signaling pathway at false discovery rate (FDR) < 5% (*P_FDR_* = 1.3 × 10^−4^) and receptor ligand activity (*P_FDR_* = 5.2 × 10^−4^) were enriched in Gene Ontology biological process and molecular function categories, respectively. Additionally, KEGG pathway analysis identified the cytokine-cytokine receptor interaction pathway (*P_FDR_* = 1.2 × 10^−3^). These results indicate that this locus is associated with multiple proteins involved in receptor ligand activity and immune defense, as indicated by IL1B and COLEC11 proteins.

**Fig. 3.**
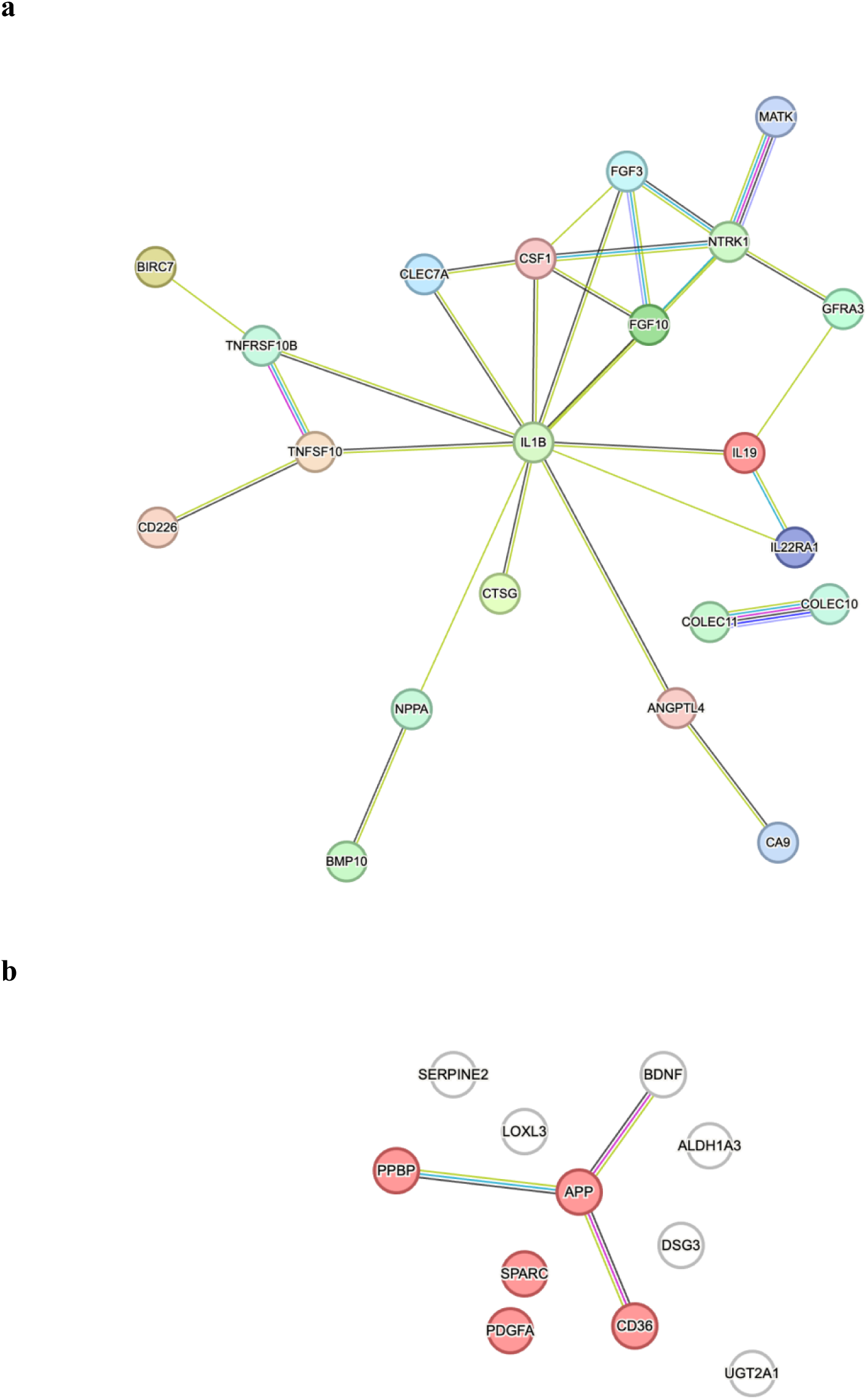
Protein-protein interaction networks of proteins associated with LD loci. a, 22 proteins are associated with the *COLEC11* gene locus. **b**, 11 proteins are associated with *CD36* gene locus. Red nodes indicate proteins involved in the platelet degranulation pathway. Protein names are listed on each node. The color of the lines indicates the type of evidence annotated using the STRING database: red, fusion evidence; green, neighborhood evidence; blue, co-occurrence evidence; purple, experimental evidence; light green, text mining evidence; light blue, curated database evidence; black, co-expression evidence; and light purple, protein homology.

We investigated 12 Japanese-specific *trans*-pQTL, comprising 10 proteins (ALDH1A3, BDNF, DSG3, PPBP, SERPINE2, SPARC, APP, PDGFA, UGT2A1, LOXL3). All 12 pQTLs were associated with a *trans*-acting variant (chr7:80569431:C:T) located in the *CD36* gene and were grouped into one locus (chr7_L30) after LD clumping. We also identified a *cis*-pQTL for the CD36 protein itself, but its lead variant (chr7:80656687:C:T) was in a separate locus (chr7_L6). Notably, this *cis*-variant was not in LD with the *trans*-variant (*r^2^* = 0.01), indicating that the *trans*-effects on the 10 other proteins are genetically distinct from the *cis*-regulation of CD36. To explore potential biological pathways underlying interactions between the CD36 protein and the other 10 proteins, we performed a Reactome pathway analysis of these 11 proteins (CD36, ALDH1A3, BDNF, DSG3, PPBP, SERPINE2, SPARC, APP, PDGFA, UGT2A1, LOXL3). This revealed a significant enrichment for the platelet degranulation pathway (*P_FDR_* = 1.2 × 10^−5^), including five proteins (CD36, APP, PPBP, SPARC, PDGFA; **Fig. 3b**). This biological link was further validated by a PheWAS search in BioBank Japan, which showed that the *trans*-variant (chr7:80569431:C:T) was strongly associated with platelet count (beta =-0.02, *P* = 2.7 × 10^−7^).

### MR study of pQTLs and diseases

To explore the potential use of proteins for causal inference, we conducted a two-sample MR study. The exposures included 561 SOMAmers associated with 714 *cis*-acting variants (**Supplementary Table 6**). For the outcomes, we selected 65 disease phenotypes, each with more than 1,000 cases from the BioBank Japan GWAS repository (**Supplementary Table 7**). Our MR analysis identified 42 putative causal associations between 26 SOMAmers (representing 24 unique proteins) and 20 distinct disease outcomes (**Table 1**). These results were validated using a colocalization test with PP.H4 > 0.8, and Steiger filtering confirmed the assumed causal direction from protein level to disease, providing no evidence of reverse causation (*P* > 0.05) (see **Methods**). We identified six cardiovascular diseases—myocardial infarction, stable angina pectoris, unstable angina pectoris, aortic aneurysm, angina pectoris, and peripheral arterial disease—that were associated with the following 11 proteins: IL6R, ADH1B, APOA5, C1QC, ABO, HP, MANBA, B3GNT8, ADH1C, PCYOX1, and MAN1C1.

**Table 1.** MR results for 20 diseases. Exposure: protein name, SeqId: SOMAmer ID, IVW: inverse variance weighted test, Wald: Wald ratio test, nsnp: number of variants used to compose instrumental variables for each protein, OR: odds ratios, PP.H4: posterior probability for colocalization test with pQTL and disease GWAS.

For diseases of the digestive system, we identified cholelithiasis, liver cirrhosis, and gastric ulcer as being associated with four proteins: FUT3, MICB, ABO, and KLB. Esophageal and hepatic cancers were linked to three proteins: ADH1B, ADH1C, and MICB. Taking FCRL1 as an example, we identified that increased FCRL1 protein levels were causally associated with an increased risk of Graves’ disease (OR =1.43, *P* = 8.67 × 10^−9^). The colocalization results indicated that the pQTL and disease GWAS signals colocalized (**Fig. 4**, PP.H4 = 0.85). Furthermore, the variant (rs7546623C>T), which was associated with FCRL1 protein levels was also associated with multiple diseases derived from the BioBank Japan PheWeb, including Graves’ disease (OR =1.17, *P* = 8.7 × 10^−9^), rheumatoid arthritis (OR =1.06, *P* = 6.2 × 10^−8^), and autoimmune disease (OR =1.10, *P* = 8.8 × 10^−7^).

**Fig. 4.**
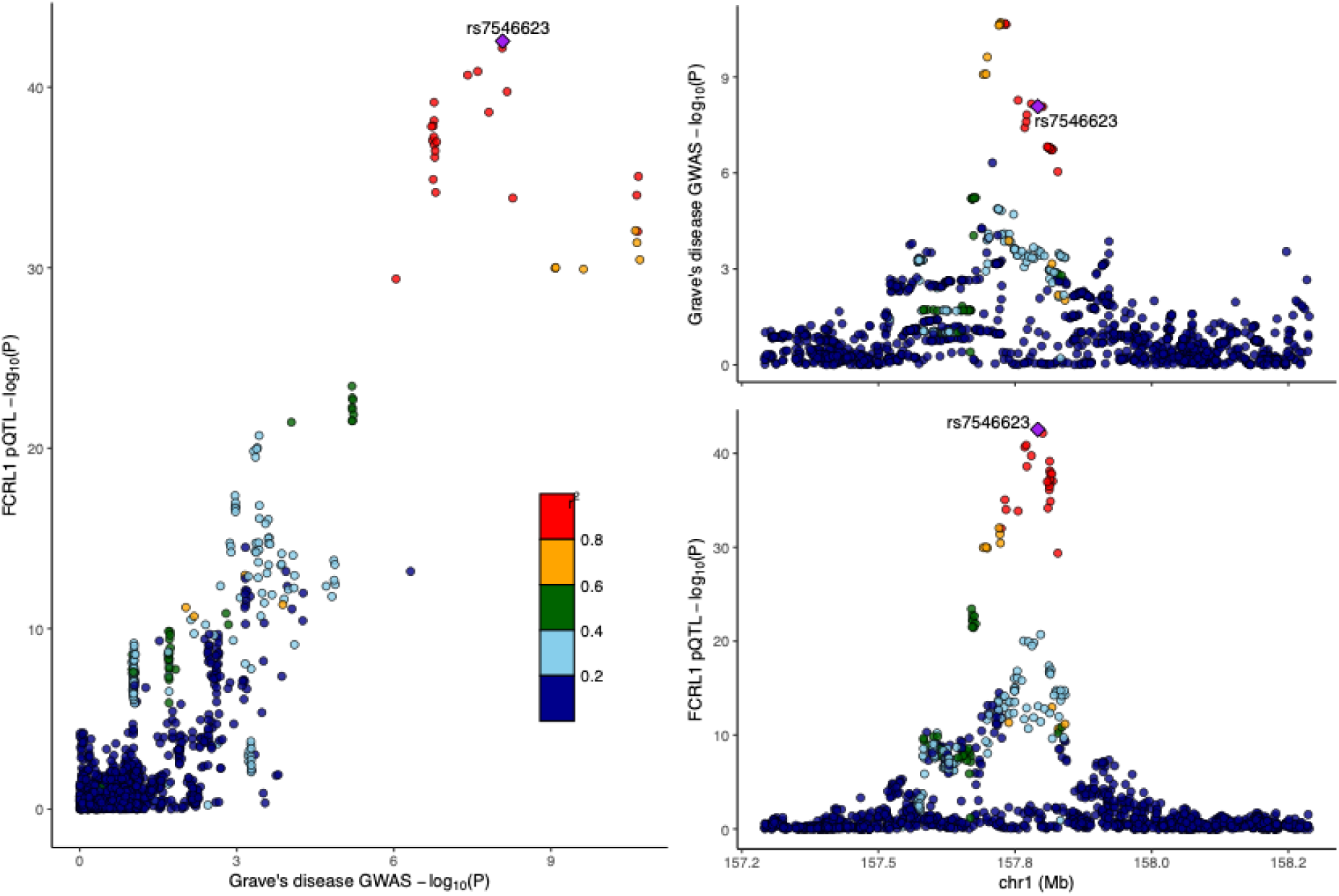
Colocalization of pQTL (FCRL1) and GWAS (Grave’s disease) The right panels display LocusZoom plots for the FCRL1 pQTL (bottom) and Graves’ disease GWAS (top). The left panel shows a scatter plot of P-values comparing the two traits. Linkage disequilibrium (*r^2^*) is indicated by color.

To identify potential therapeutic targets, we queried 24 proteins using ChEMBL (ChEMBLdb version 35). Through this approach, we identified 11 proteins targeted by 116 drugs that have either been approved or are currently undergoing clinical trials (**Table 2** and **Supplementary Table 8**). Of the 116 drugs, 41 targeted IL6R, 40 targeted HP, and 13 targeted IL1RL1 or IL18R1. MR analysis identified IL18R1 and IL1RL1 as causal proteins for atopic dermatitis. Among the 13 drugs targeting IL18R1 and IL1RL1, astegolimab and spesolimab were identified as treatments for atopic dermatitis (**Supplementary Table 8**). Furthermore, we also identified an inhibitor of FCRL1, obexelimab, which is currently in phase 3 clinical trials for IgG4-related disease.

**Table 2.** Potential therapeutic targets for drug repurposing. Drug Name: the name of the drug, Action Type: type of action the drug has on the protein, Mesh: Medical Subject Headings.

## Discussion

This extensive proteogenomics study represents the most comprehensive pQTL analysis conducted to date for large-scale protein quantification within the Japanese population. Based on 4,392 SOMAmers and 4.6 million variants measured in 1,823 individuals, we identified 1,876 associations, including 77 newly identified pQTLs specific to the Japanese population. Our study demonstrates that integrating plasma protein levels, expression datasets, and disease phenotypes can enhance our understanding of the plasma proteome. This approach clarifies how population-specific variants influence both protein levels and clinical phenotypes. For example, we identified two Japanese-specific variants in the *CD36* gene that were associated with 12 proteins and with decreased white blood cell and platelet counts.

Our multi-ancestry fine-mapping and colocalization analyses provide insights into the genetic architecture linking gene expression with plasma protein levels. We found that 170 of 656 measured proteins (26%) shared *cis*-pQTL credible sets between individuals of Japanese and European populations. However, when colocalizing these *cis*-pQTLs with *cis*-eQTLs from the Nagahama whole blood dataset, we identified shared causal variants for only 72 proteins (11%). This colocalization rate is notably lower than proportions reported in the GTEx dataset. This discrepancy may be attributed to the tissue context, as the Nagahama dataset was generated from whole blood. The use of whole blood gene expression represents a limitation in our study, as it fails to capture tissue-specific regulatory signals. This is particularly relevant given that 411 of the 656 genes encoding the measured proteins are expressed in a tissue-specific manner. These results indicate the complex interplay of genetic background, tissue-specific regulation, protein subcellular location, and the potential effects of PAVs on protein detection.

The combined analysis of *cis*- and *trans*-pQTL revealed possible protein interactions within specific pathways, exemplified by the locus near *COLEC11,* which was associated with 46 SOMAmers. Among these 46 proteins, we observed enrichment of the cytokine-cytokine receptor interaction pathway, with IL1B and COLEC11 participating in immune defense. Furthermore, we also identified a Japanese-specific *trans*-pQTL hotspot located in the *CD36* gene, which was associated not only with 10 proteins but also with other phenotypes, such as lower platelet and white blood cell counts in plasma. Therefore, by combining the results of *cis*- and *trans*-pQTLs, we identified pleiotropic loci and elucidated key biological pathways connecting these associated proteins.

Finally, our MR analysis integrating *cis*-pQTLs revealed 42 putative causal protein-disease associations and highlighted potential opportunities for clinical translation. Specifically, by querying 24 proteins from the MR results, we identified 11 potential drug targets associated with 116 drugs. The fact that 53 of these drugs are already approved underscores the substantial potential for drug repurposing. This analysis demonstrates a clear pathway from genetic association to clinical translation, helping to identify novel biomarkers and prioritize actionable druggable targets.

Despite our efforts, this study has certain limitations. First, the total number of variants used for WGS analysis was smaller than that used in some WGS studies, owing to our stringent quality control criteria. These criteria were set to obtain high-quality variants from two variant-calling pipelines with varying sample sizes and to eliminate any possible bias in the results. Consequently, approximately 12.5 million variants were removed. This issue can be addressed by employing a single variant-calling pipeline in the future and using a reference panel from a larger Japanese cohort to enhance sensitivity for detecting rare variants.

Second, while multi-ancestry fine-mapping analyses revealed high correlations in *cis*-pQTL effect sizes between Japanese and European populations, a larger, tissue-specific East Asian gene expression dataset is still needed. Such a dataset would enhance the statistical power of colocalization analyses and help to further elucidate the shared genetic etiology between gene expression and plasma protein levels. Lastly, we excluded *trans*-pQTLs from our MR analysis to minimize horizontal pleiotropy, as these variants can affect the outcome via pathways unrelated to the target protein. This decision resulted in instruments composed of only a small number of *cis*-acting variants. Therefore, our approach reduces the risk of pleiotropy but also limits the explained variance, thereby decreasing statistical power. While including *trans*-pQTLs would have improved power and permitted more comprehensive sensitivity analyses, such as the MR-Egger intercept test or other outlier-robust methods, including MR-PRESSO^39^ and weighted median^40^ methods, it would also require additional assessment, as *trans*-acting variants may affect outcomes through pathways unrelated to the target protein.

In conclusion, our findings provide significant insights into the genetic regulation of the human plasma proteome within the Japanese population. We highlight how population-specific variants may influence disease pathogenesis through the plasma proteome, reinforcing the need to conduct proteogenomic studies across diverse ancestries. The data generated by this study serve as a valuable resource for deepening the understanding of disease mechanisms and facilitating targeted drug development.

## Code availability

R v4.1.0: https://www.r-project.org/

PLINK v2.00a3LM: https://www.cog-genomics.org/plink/2.0/

GCTA-COJO v1.94.1: https://yanglab.westlake.edu.cn/software/gcta/#COJO

coloc v5.1.0: https://chr1swallace.github.io/coloc/

MESuSiE: https://github.com/borangao/MESuSiE

TwoSampleMR v.0.5.6: https://mrcieu.github.io/TwoSampleMR/

## Data availability

Nagahama *cis*-pQTL summary statistics: https://www.hgvd.genome.med.kyoto-u.ac.jp/repository/HGV0000026.html

(Full pQTL summary statistics will be made available upon acceptance.) Nagahama *cis*-eQTL summary statistic (HGVD eQTL v8.1): https://www.hgvd.genome.med.kyoto-u.ac.jp/download/eQTL/

ARIC summary statistics (European and African-American): http://nilanjanchatterjeelab.org/pwas/

deCODE summary statistics: https://www.deCODE.com/summarydata/

GTEx eQTL v8: https://www.gtexportal.org/home/downloads/adult-gtex/qtl

BioBank Japan disease GWAS (Sakaue, S. & Kanai, M., et al.): https://pheweb.jp/downloads

## Supporting information

Main Table 1 and 2

## Data Availability

All data produced in the present work are contained in the manuscript

https://www.hgvd.genome.med.kyoto-u.ac.jp/repository/HGV0000026.html

## Acknowledgments

We thank Miki Kokubo for technical assistance in this study. We are grateful to the Nagahama City Office and the non-profit organization Zeroji Club for their assistance in conducting the Nagahama study. We thank Chen-Yang Su for the critical review of the manuscript. The study was partly supported by the Practical Research Project for Rare/Intractable Diseases of the Ministry of Health, Labour and Welfare (2011-Ip-pan-002) and Japan Agency for Medical Research and Development (JP23ek0109675, JP17ek0109283, JP14ek0109070) and operational funds of Kyoto University for the Top Global University Project and Takeda Science Foundation.

## Author contributions

Ta-Yu Yang (T.-Y.Y.), Masao Nagasaki (M.N.), and Fumihiko Matsuda (F.M.) designed the study. Takahisa Kawaguchi (T.K.), Kazuya Setoh (K.S.), Yasuharu Tabara (Y.T.), Takeo Nakayama (T.N.), and Shinji Kosugi (S.K.) participated in the collection of biological materials and clinical information from participants, along with the related analysis of the Nagahama Study. Katsunori Horii (K.H.), Shintaro Kato (S.K.), and Iwao Waga (I.W.) conducted plasma protein measurements. Meiko Takahashi (M.T.) conducted and supervised the WGS experiment. Koichiro Higasa (K.H.), Takahisa Kawaguchi (T.K.), and Masao Nagasaki (M.N.) conducted quality control and analyzed the WGS data. Zhao Tie (Z.T.) analyzed the Nagahama eQTL. Ta-Yu Yang (T.-Y.Y.) and Takahisa Kawaguchi (T.K.) conducted proteomic and statistical analyses. Satoshi Yoshiji (S.Y.) and Takeshi Iwasaki (T.I.) provided key comments and methods during the pQTL analyses. Ta-Yu Yang (T.-Y.Y.), Masao Nagasaki (M.N.), and Fumihiko Matsuda (F.M.) drafted the manuscript. Shuji Kawaguchi (S.K.), Satoshi Yoshiji (S.Y.), Takeshi Iwasaki (T.I.), Shintaro Kato (S.K.), Kazuya Setoh (K.S.), Masao Nagasaki (M.N.), and Fumihiko Matsuda (F.M.) reviewed the paper and provided key comments. All authors have approved the final version of the manuscript.

