## Supplemental Figures 1 to 8 for "Comprehensive genomic atlas of plasma proteome in the Japanese population: the Nagahama study"

### Supplementary Figures

Supplementary Figure 1. Correlation of effect sizes between Japanese and European populations.

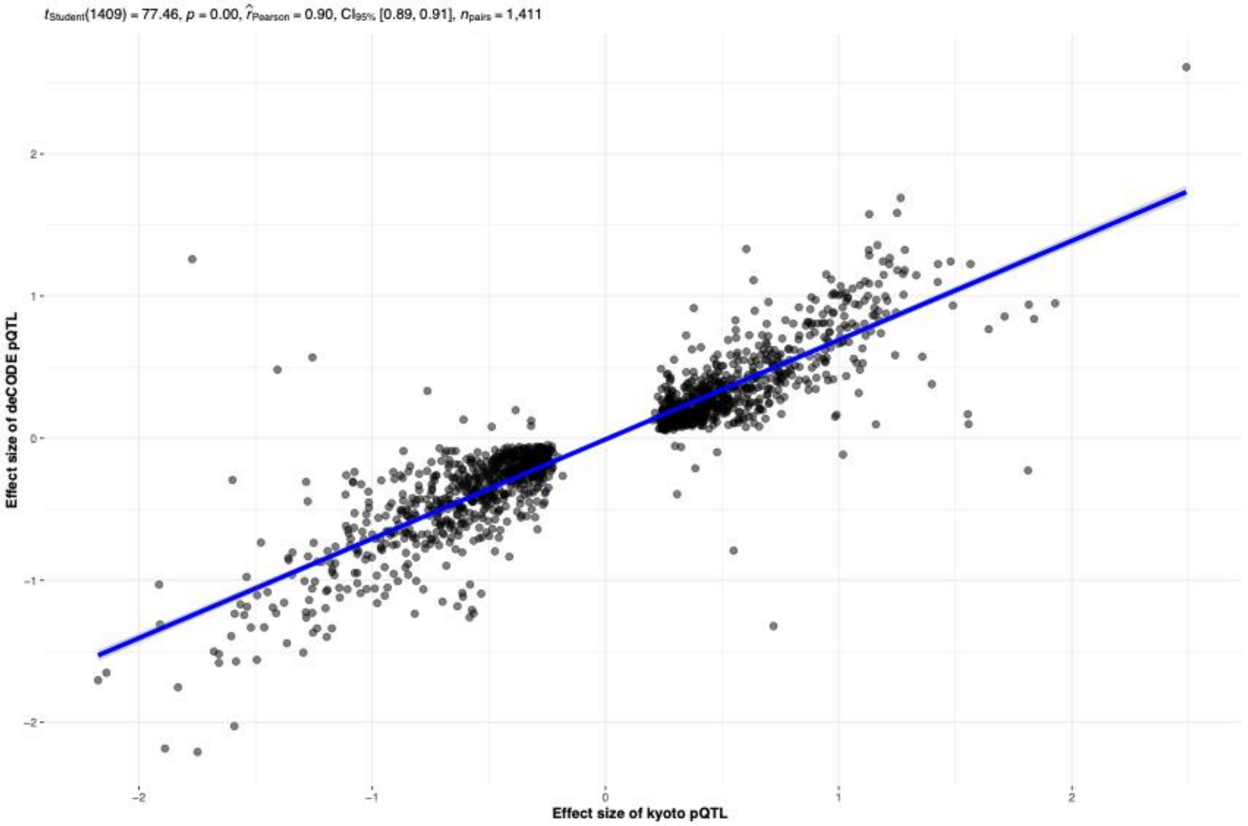

Supplementary Figure 2. Correlation of effect sizes between Japanese and African American populations.

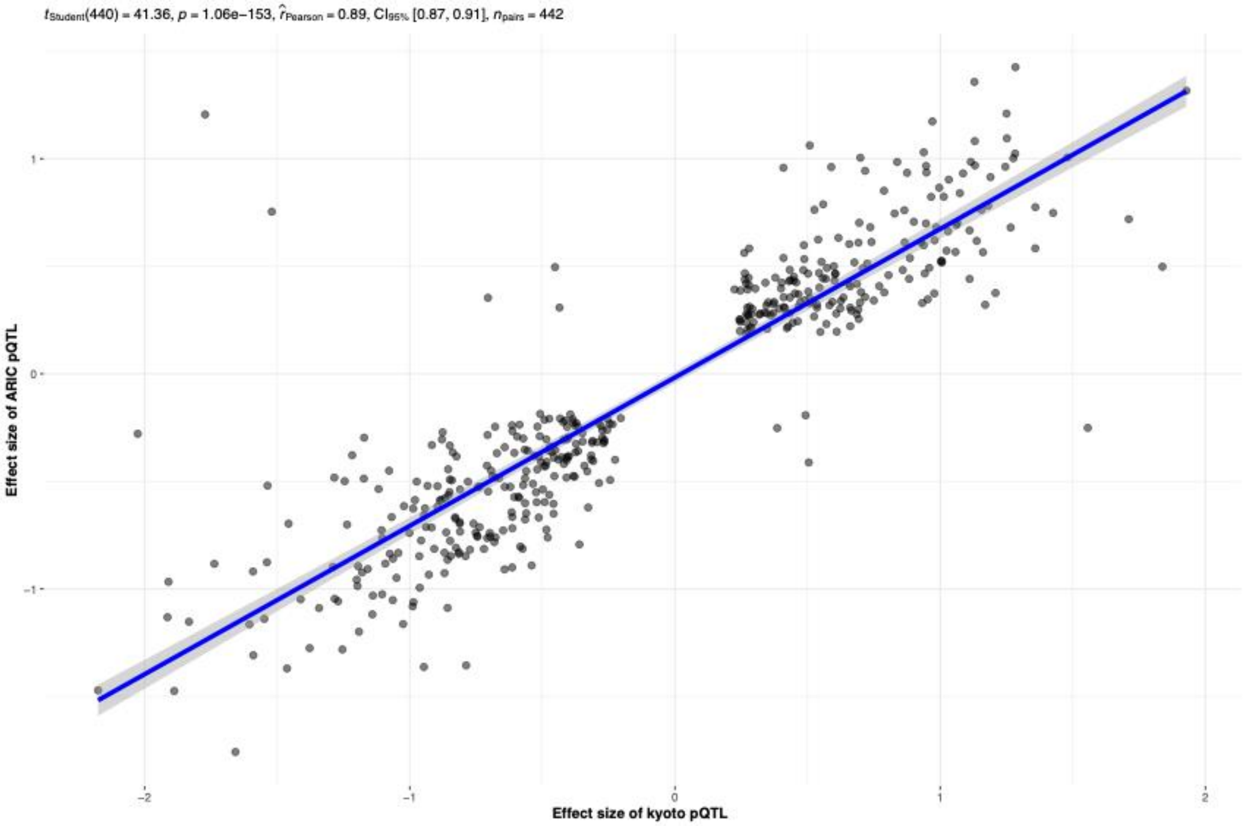

Supplementary Figure 3. Correlation of effect sizes between *cis*-pQTLs and *cis*-eQTLs (GTEx).

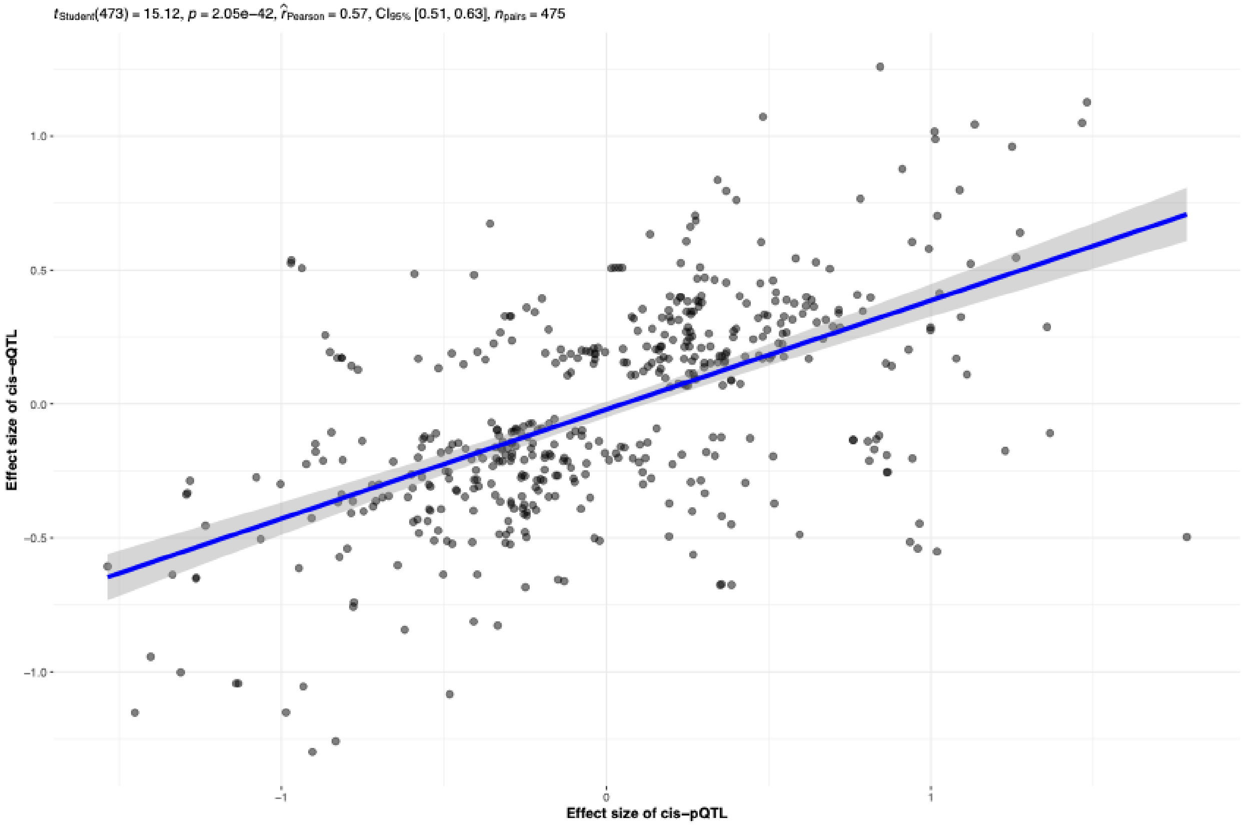

Supplementary Figure 4. Correlation of effect sizes between *cis*-pQTLs (secreted or membrane proteins) and *cis*-eQTLs (GTEx).

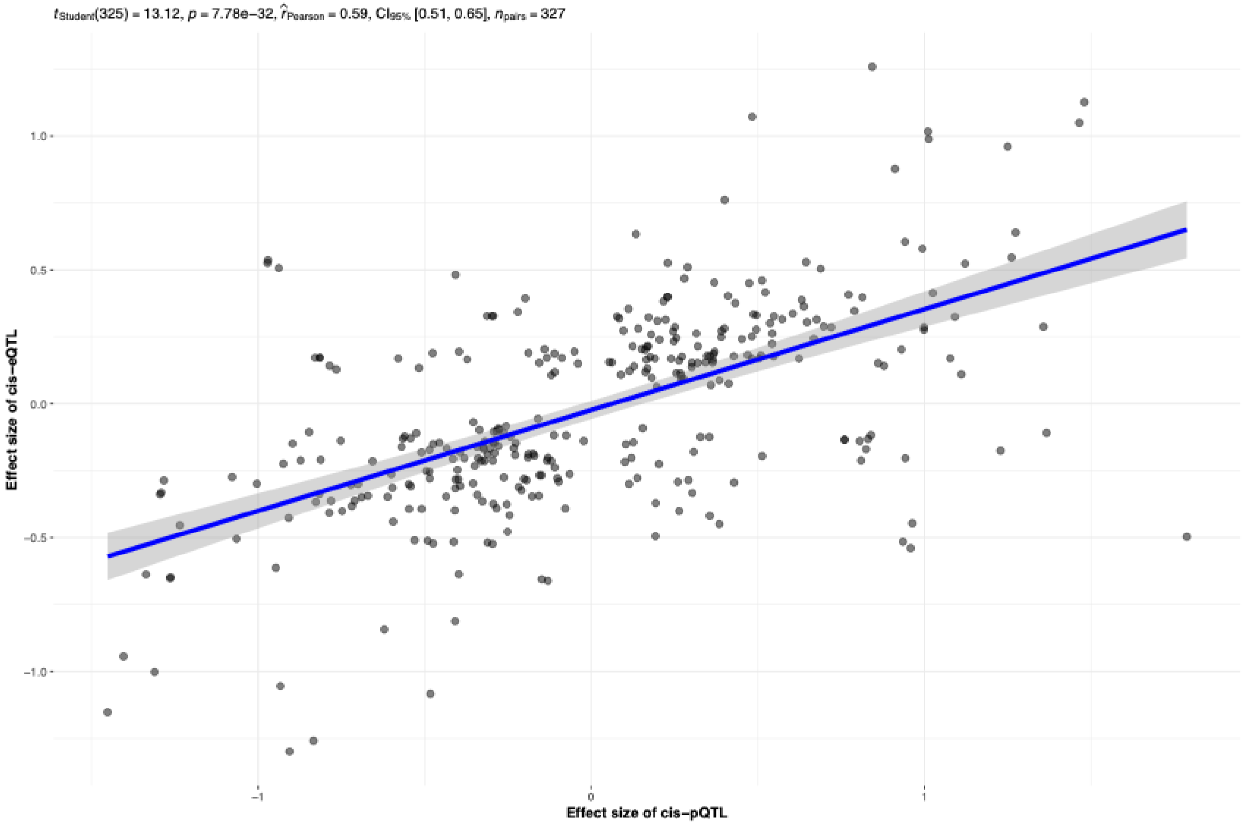

Supplementary Figure 5. Correlation of effect sizes between *cis*-pQTLs (intracellular proteins) and *cis*-eQTLs (GTEx).

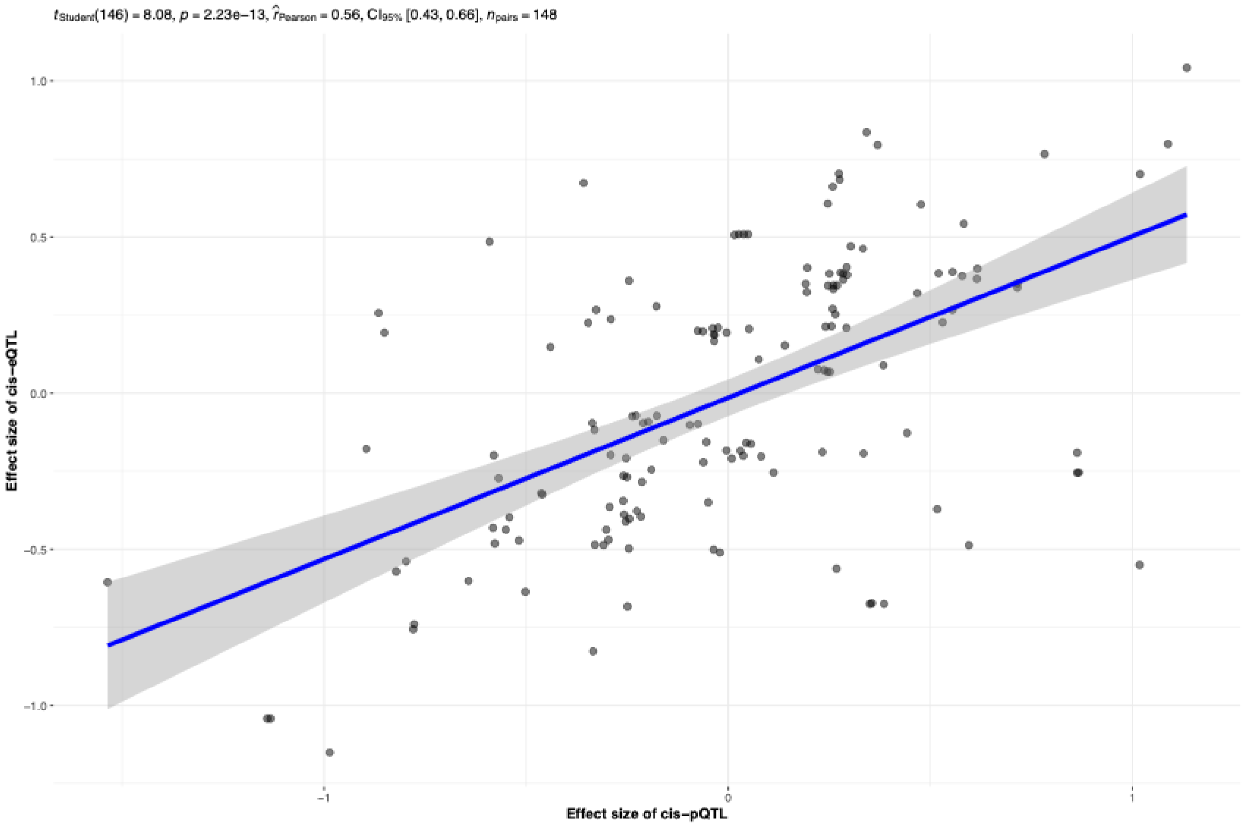

Supplementary Figure 6. Correlation of effect sizes between *cis*-pQTLs (secreted or membrane proteins; PAVs) and *cis*-eQTLs (GTEx).

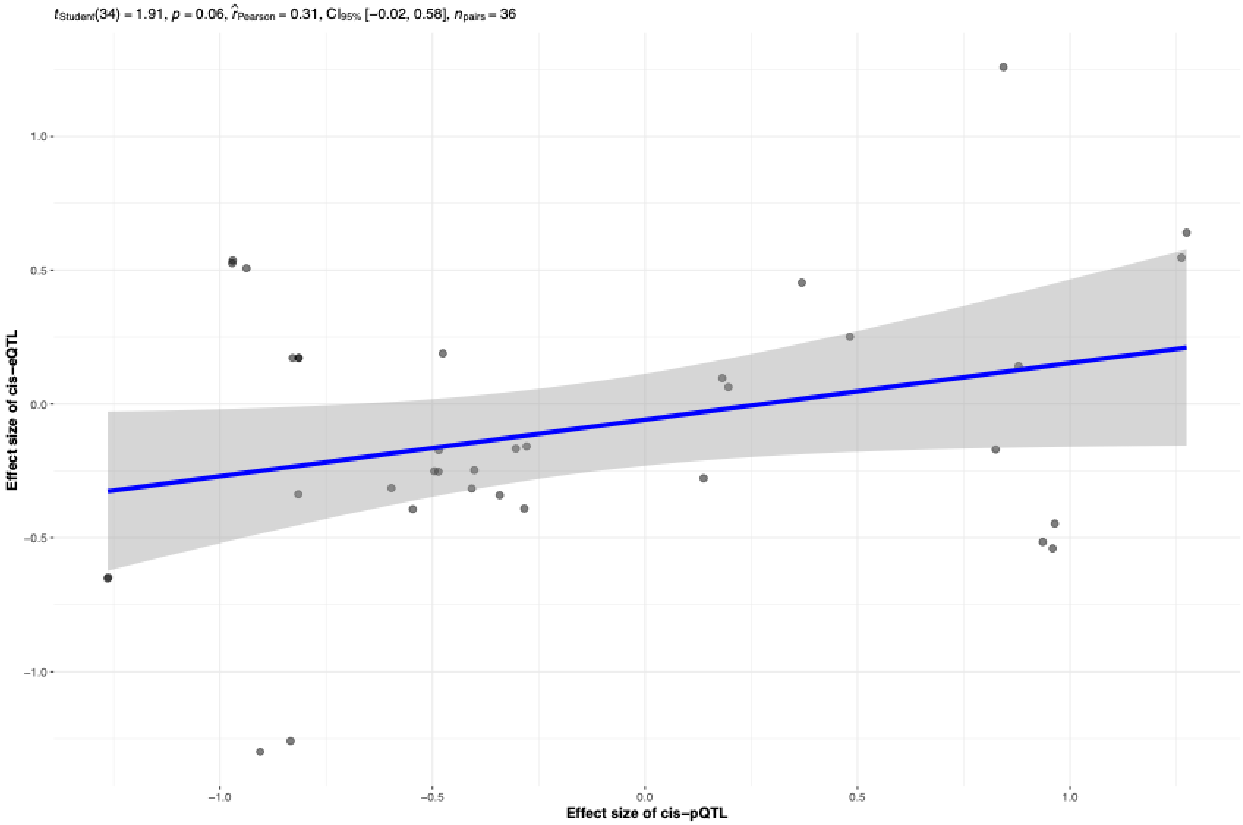

Supplementary Figure 7. Correlation of effect sizes between *cis*-pQTLs (secreted or membrane proteins; non-PAVs) and *cis*-eQTLs (GTEx).

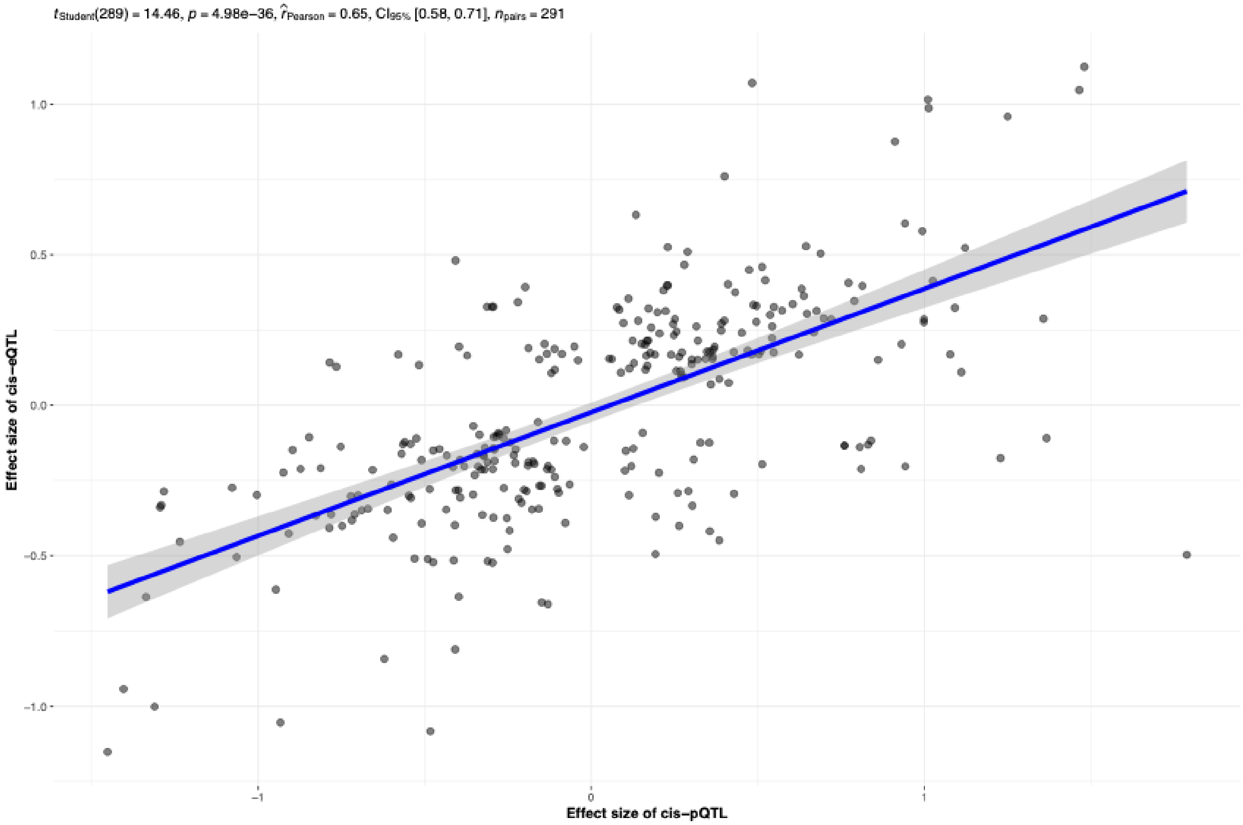

Supplementary Figure 8. Correlation of effect sizes between Nagahama *cis*-pQTLs and Nagahama *cis*-eQTLs (Whole blood).

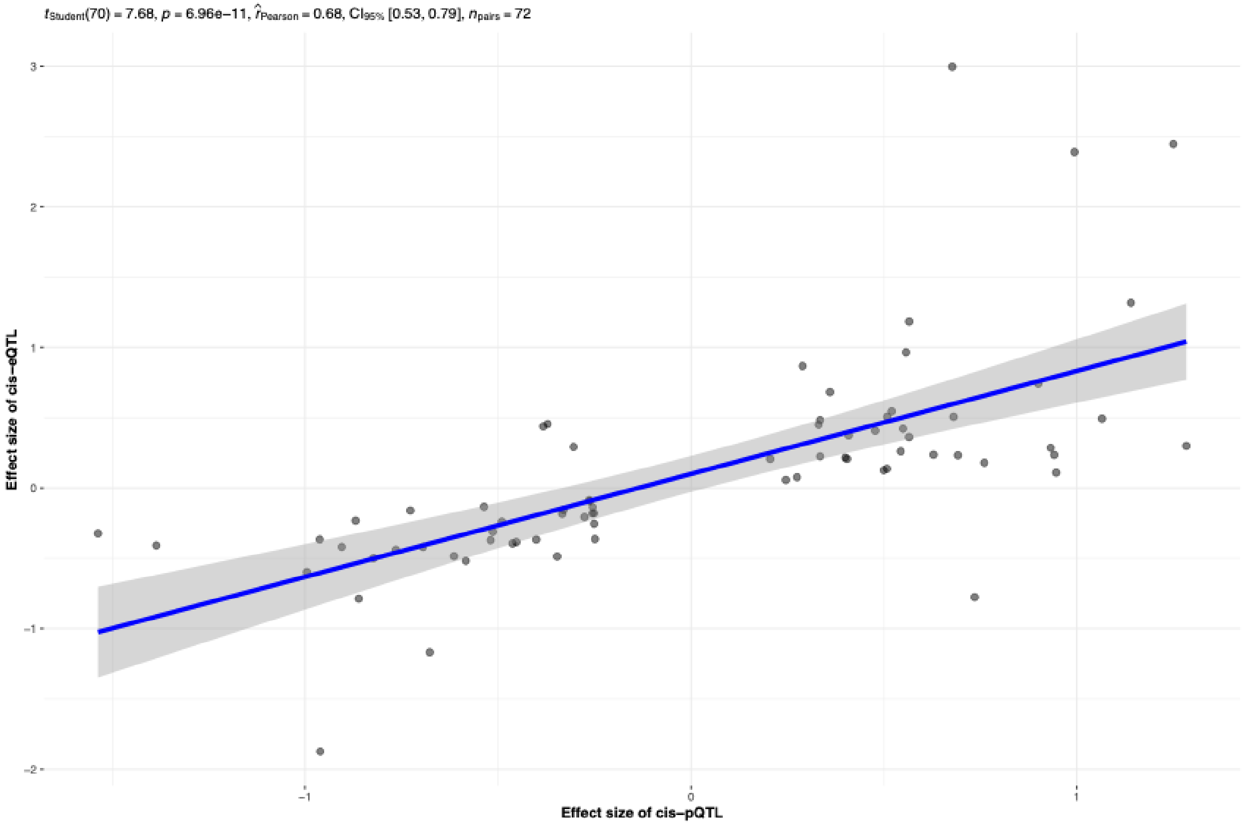
